# Modelling the Excess Mortality Associated with Heat Waves in Hong Kong: 2014-2023

**DOI:** 10.64898/2026.03.05.26347683

**Authors:** Zhenyuan Liu, Chao Ren, Jingwen Liu, Kawasaki Yurika, David Makram Bishai

## Abstract

**Introduction:** Heat waves are increasingly frequent and linked to higher mortality risks in Hong Kong. However, estimates of total excess mortality associated with heat waves remain unavailable. This study quantifies excess deaths associated with heat waves in Hong Kong from 2014 to 2023.

**Methods:** Daily age- and sex-specific mortality rates and population data were obtained from the Hong Kong Life Tables and Census and Statistics Department. Temperature data came from the Hong Kong Observatory, and relative risks were derived from local research. A Monte Carlo simulation was used to estimate heat-attributable deaths under different heat wave definitions, calculating total excess deaths and annualized death rates per 100,000 population.

**Results:** Between 2014 and 2023, heat exposure resulted in an estimated 1,455 (95% CI: 1,098–1,812) to 3,238 (95% CI: 3,234–3,242) excess deaths. In 2023, annualized excess death rates ranged from 2.95 (95% CI: 2.41–3.50) to 5.09 (95% CI: 5.07–5.12) per 100,000 people. Males and individuals aged 65 or older were disproportionately affected.

**Conclusion:** Over the 10-year study period, 1,455 to 3,238 excess deaths in Hong Kong were attributed to extreme heat. Heat waves now rank among the top ten causes of death in Hong Kong, with mortality rates comparable to diabetes. These findings underscore the need for urgent public health interventions to mitigate the impact of extreme heat.

**Author Summary:** *Why was this study done?:*

- Hong Kong has experienced increasingly frequent and intense heatwaves in recent years, yet the health impacts of extreme heat have not been comprehensively quantified.
- There is a lack of localized evidence on the mortality burden attributable to extreme heat, particularly across different demographic groups, to inform public health responses in subtropical urban settings.

*What did the researchers do and find?:*

- This study modelled the association between extreme heat and all-cause mortality in Hong Kong from 2014 to 2023, applying multiple heatwave definitions.
- The results estimate that over 1,400 excess deaths could have been preventable during the study period, with elevated vulnerability observed among males and individuals aged 65 years and older.
- Extreme heat was identified as an independent risk factor for mortality, with a health burden comparable to that of chronic diseases such as diabetes.

*What do these findings mean?:*

- The findings underscore the urgent need for Hong Kong to develop and implement comprehensive Heat Action Plans (HAPs) to reduce excess mortality during extreme heat events.
- With climate change expected to increase the frequency and severity of heatwaves—and an aging population—HAPs must prioritize the protection of high-risk groups.
- These results provide critical evidence for public health planning, emphasizing the importance of tailored, data-driven interventions to enhance climate resilience in urban populations.

## Introduction

The intensification of climate change in recent years has resulted in more frequent heatwaves.^1^ According to a recent World Meteorological report, there is an 80 percent likelihood that the annual average global temperature will temporarily exceed 1.5°C above pre-industrial levels for at least one of the next five years,^2^ making exposure to extreme heat a growing public health concern. Extreme heat disrupts the body’s ability to regulate temperature, leading to serious health consequences and exacerbating chronic conditions, which increases mortality risk that were well-documented by previous studies.^3, 4, 5^ Furthermore, the adverse effects are exacerbated in cities with high population density and outmoded urban architecture, such as poorly insulated buildings and inadequate ventilation systems, where urban heat islands further intensify indoor temperatures during heat waves.^6, 7^

There is now compelling evidence to demonstrate the general relationship between temperature and mortality, as observed in various settings worldwide, including Hong Kong. These studies use daily time series data on deaths and temperatures and estimate the probability of dying on or after a hot day in terms of relative risks,^8, 9, 10, 11^ attributable fractions,^9^ and excess mortality rate.^12, 13^ However, absolute numbers of how many people died are more effective than relative risks in conveying the total burden of mortality during extreme heat events to the public and stakeholders.^14^ Some studies have reported the number of heat-attributable deaths,^9, 15^ but attributing deaths solely to heat is complex and may underestimate heat-related mortality, as heat exposure can exacerbate pre-existing medical conditions, thereby increasing deaths from causes beyond heatstroke and hyperthermia.^16^ Fewer studies estimate excess deaths during heatwaves—calculated as the difference between observed and expected deaths under normal conditions—an approach that mitigates ascertainment bias from death records that often fail to attribute heat as the underlying cause.^17, 18, 3^ Given the above, excess death measures are advantageous due to their objectivity, replicability, and high face validity.

To quantify the excess mortality associated with heatwaves and to effectively communicate the health implications of prolonged elevated temperatures, we utilized published estimates of relative risk (RRs) of mortality during heat events specific to Hong Kong.^9,11^ By estimating the absolute number of excess deaths and the age-standardized excess death rate per 100,000 population, we provide a critical metric for assessing the mortality burden associated with heat exposure.

## Method

### Definition of heat waves

Given the absence of a universally accepted definition for a heat wave, we employed four distinct definitions that capture heat wave conditions in Hong Kong, as delineated in previous local studies (**Table 1**). The first definition identifies a heat wave as a 20-day period commencing when the daily mean temperature exceeds the 99th percentile threshold of 30.60 °C. This duration accounts for the lagged effects of a single hot day, which may persist for up to 20 days, as estimated by Liu et al. (2020).^9^ Additionally, to qualify as an independent heat wave event, the initial day must be separated by at least 20 days from the preceding day that surpassed the threshold. The second definition characterizes a heat wave based on the number of consecutive days with daily maximum temperatures exceeding 33 °C. The third definition focuses on the number of consecutive days with daily minimum temperatures above 28 °C. Lastly, the fourth definition determines a heat wave by the simultaneous occurrence of hot days and hot nights, following the metrics established by Wang et al. (2019).^11^ These diverse criteria were selected for our analysis based on their well-established associations with increased excess mortality risks, while controlling for lagged effects and common confounding factors such as humidity, air pollutants, and seasonality.

**Table 1.**
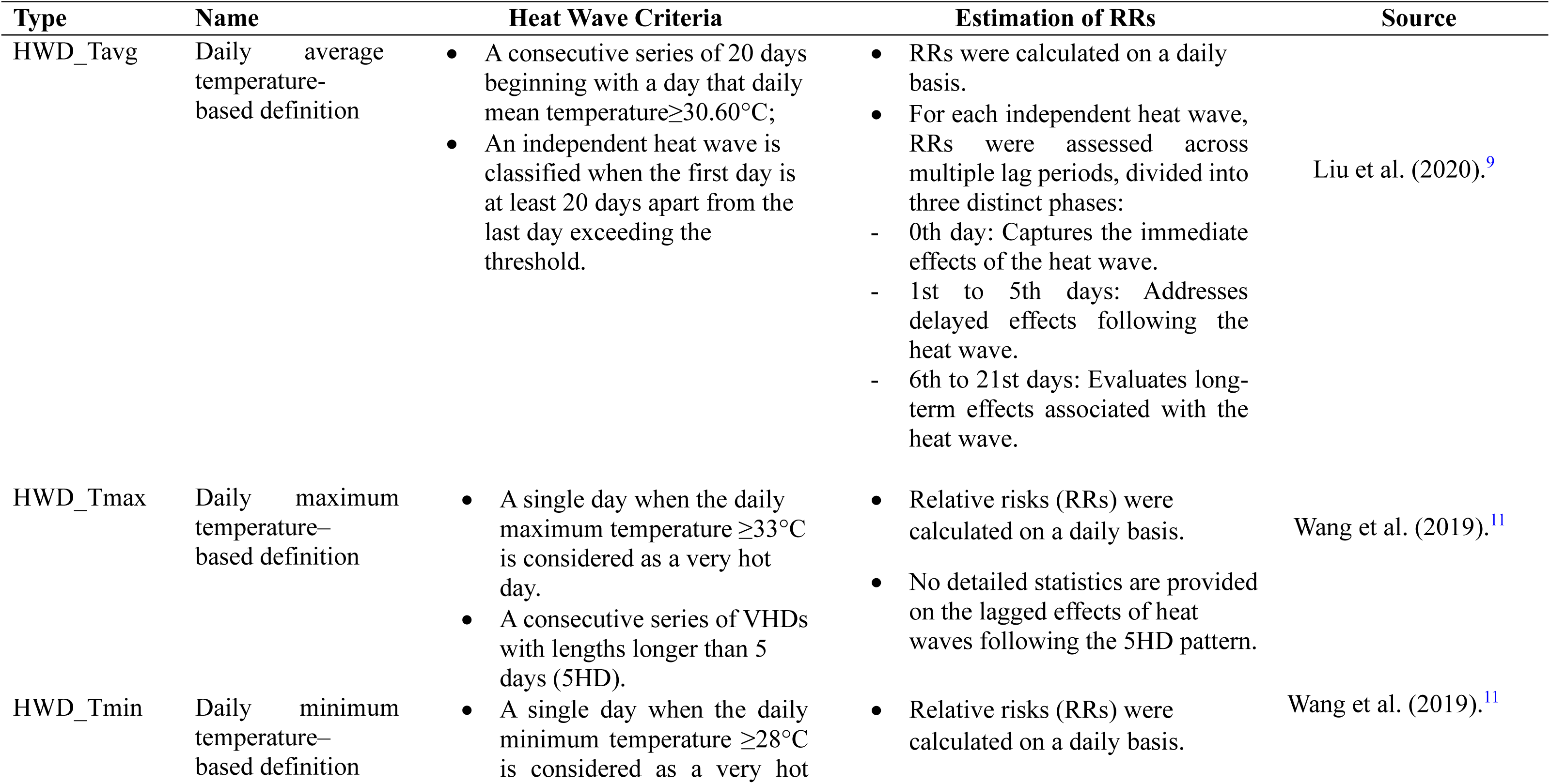
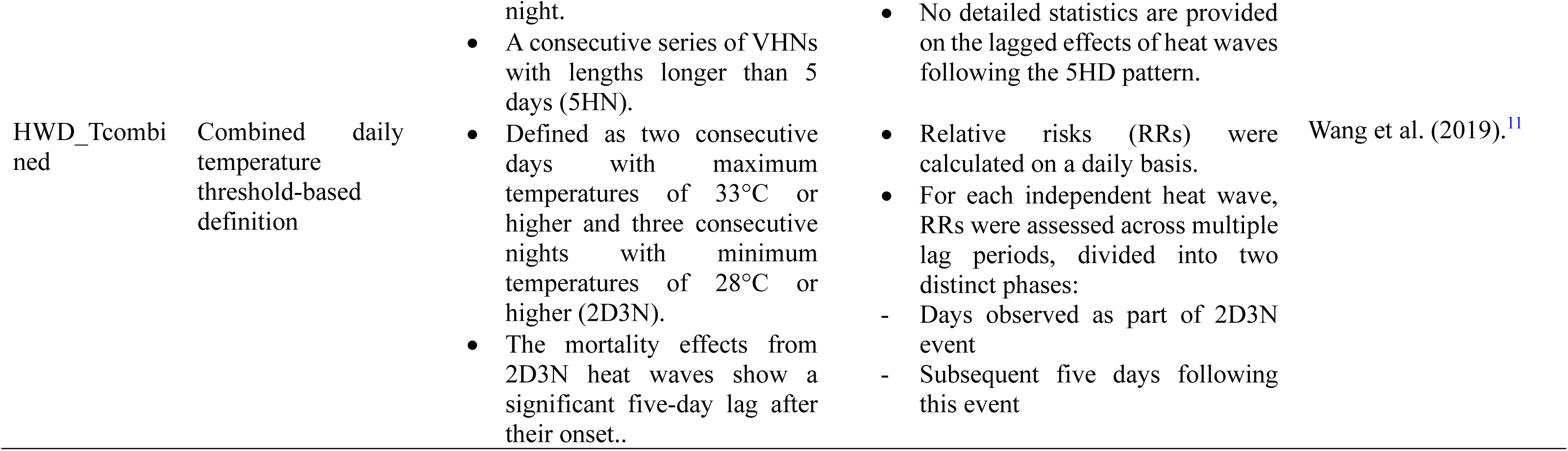
Heat wave definitions used as the basis for the present analysis.

Daily mean temperatures from 2014 to 2023 were obtained from the Hong Kong Observatory (HKO) station,^19^ chosen for its comprehensive data coverage. This dataset enabled us to identify and characterize heat wave events under the four different scenarios over the past decade.

### Daily number of deaths

A key component of calculating excess heat-related deaths (EHD) is the deaths that occur under normal conditions (no heat wave). Expected daily mortality data, disaggregated by age-sex specific group, were sourced from the Hong Kong Life Tables covering the period from January 1, 2014, to December 31, 2023.^20, 21^ Subsequently, a time series of normal death counts was generated by multiplying the age-sex specific death rates by the corresponding population figures from the Hong Kong Census and Statistics Department.^22^

### Transformation of relative risks

We used estimates of the RRs due to heat from Liu et al. (2020)^9^ and Wang et al. (2019).^11^ The theoretical basis for EHD involves a counterfactual comparison, contrasting observed conditions (e.g., heat exposure) with a hypothetical reference state (e.g., normal conditions without heat exposure). RR represents the ratio of the probability of death under heat exposure to that in a reference group under ordinary temperatures, as a key variable in calculating EHDs. All RRs are estimated at the level of the whole population, sex and age groups. Therefore, to make these RRs fit in the age-sex specific structure of Hong Kong Life Tables, ^20, 21^ we adjusted them by combining the log-transformed relative risks for age and sex while accounting for the overall population risk. Mathematically, this is represented as **Equation 1**:

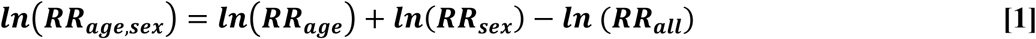

Subsequently, adjusted log-RRs are exponentiated, then reverted to the original RRs scale. Detailed adjusted RRs are provided in **Supplementary Table S1**.

Adjusted RRs and their corresponding standard deviations are also logarithmically transformed. This transformation is essential as the RRs represent a ratio, and confidence intervals for their parameters are commonly estimated on the logarithmic scale, indicating a log-normal distribution.^24^ Consequently, we generated 1000 values of log-normally distributed RRs, using a pseudo-random number generator in Stata by using **Equation 2** as follows:

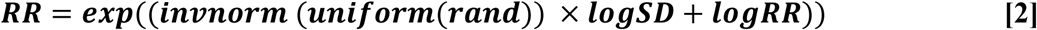

### Estimation of excess mortality attributable to extreme heat

We subsequently calculated the EHDs for each single heat wave observed in the 10-year interval. The expected number of excess deaths attributable to extreme heat over various lag days for the t-th heat wave was determined using **Equation 3**: ^25, 26^

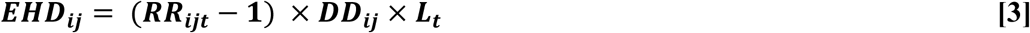

Specifically, ***i*** denotes sex, ***j*** denotes age group. ***L***_***t***_ signifies the duration in days of the t-th phase of a heat wave. ***RR***_***ijt***_ is the relative risk of deaths for sex i, age group j, during the t-th phase of a heat wave, while ***DD***_***ij***_ is the expected number of daily deaths in the counterfactual scenario of no heat wave. We generated a distribution of 1000 random estimates of the RRs using **Equation 2**, then drew from the distribution to estimate ***EHD***_***ij***_ using Monte Carlo simulation. The simulated results are expressed as ***EHD***_***ijn***_, where n ranges from 1 to 1000, representing the Monte Carlo simulation index. This method is detailed in Zhao et al. (2024).^27^

Finally, we multiplied the cumulative count of heat waves in Hong Kong for each year from 2014 to 2023 by ***EHD***_***ij***_ to get estimates of annual excess heat-related deaths which could be added up to the total deaths by age and sex in a decade. We also reported the excess mortality as age-standardized excess death rates per 100,000 population. The age adjustment was based on the world standard population as specified in the revised age-standardization of rates by the World Health Organization - 2001 Edition.^28^

The estimated excess deaths count and the age-standardized death rates across the last decade were accompanied by 95% confidence intervals (CIs). These CIs were obtained using the normal approximation with the bootstrap method described by Pizzato et al. (2024),^29^ employing 1000 samples generated through Monte Carlo simulation. Given that discounting can substantially affect mortality estimates, as seen in the Global Burden of Disease approach,^30^ we present excess heat-related deaths and death rates without discounting or age weighting, treating all deaths equally irrespective of age or when they occur.

We used Stata SE (v.18.0) for all statistical analyses. All data used in this analysis were aggregated statistics at the population level. The study was not human subject’s research.

## Results

### Summary statistics

During the study period in Hong Kong, average daily temperature was 24.12°C, ranging from 12°C to 31.1°C (**Supplementary Table S2**). As shown in Figure 1 different heat wave definitions differ in frequency and in total days of exposure over a decade. The HWD_Tcombined definition racked up the highest exposure at 365 days over 10 years, and the highest frequency of occurrences at 38 events over 10 years. The definition labeled HWD_Tavg occurred on 360 heat wave days but had the lowest frequency of 18 events. In contrast, HWD_Tmax recorded the fewest heat wave days at 183 and a frequency of 25 events. HWD_Tmin demonstrated a slightly higher frequency of 26 occurrences whereas it also had a limited duration of 190 heat wave days **(Figure 1**).

**Figure 1.**
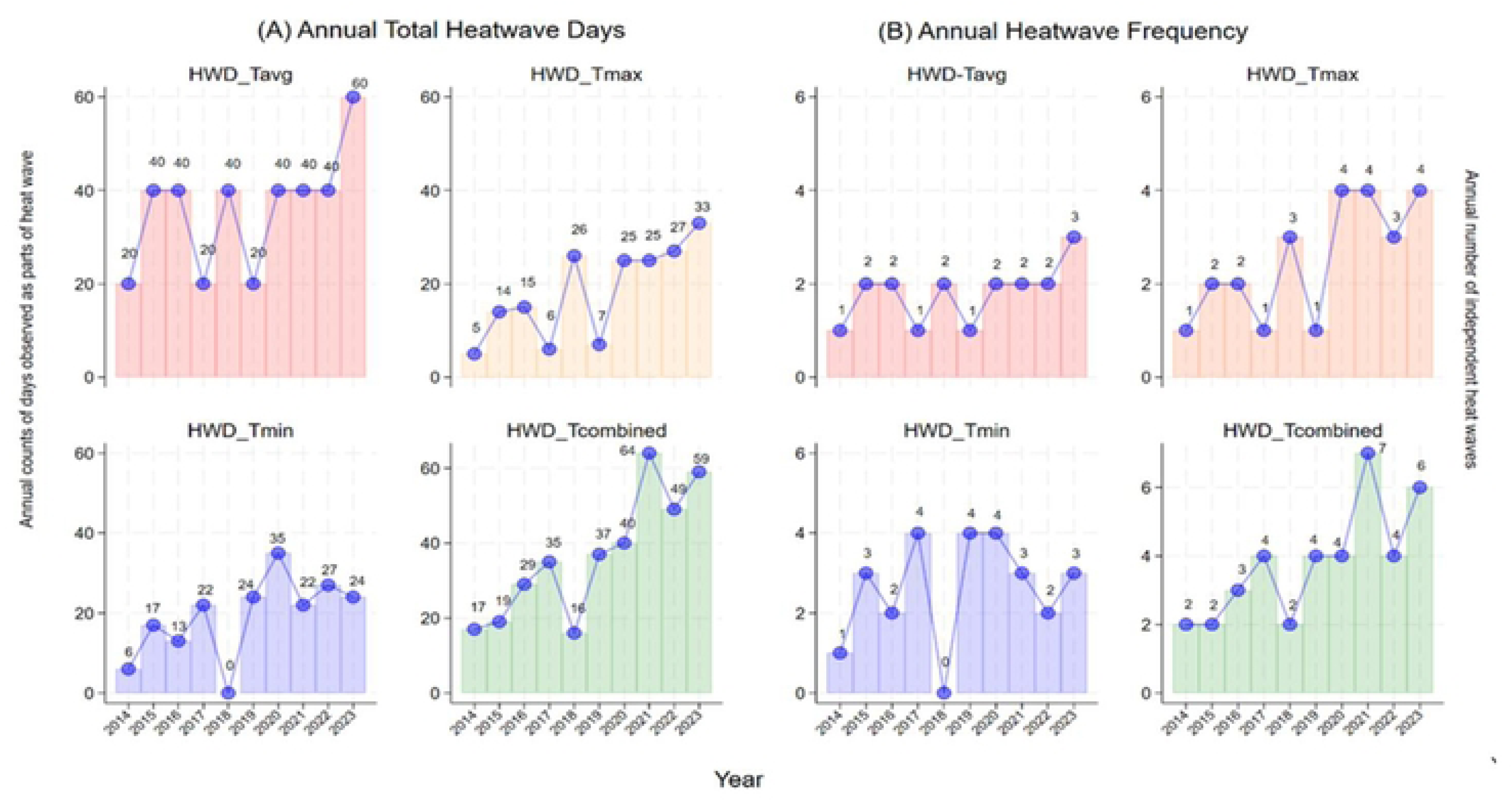
The descriptive statistics of heatwaves. A: Annual sum of frequency of heatwaves in Hong Kong, during 2014–2023; B: Annual counts of heat wave days in Hong Kong, during 2014–2023.

### Excess Mortality in number and age-standardized death rate

Based on the definition of heat event, the estimates of total excess deaths attributable to heat waves ranged from 1,455 (95% CI: 1,098–1,812) to 3,238 (95% CI: 3,234–3,242) (**Table 2** and **Figure 2A**). Notably, the heat wave definition based on combined temperature threshold metrics (HWD_Tcombined) resulted in more than double the estimated excess deaths compared to estimates derived from mean daily temperature (HWD_Tavg) (**Table 2**). The HWD_Tavg definition gave the lowest cumulative estimates of excess heat-related deaths. See **Table 2** and **Figure 2A**.

**Figure 2.**
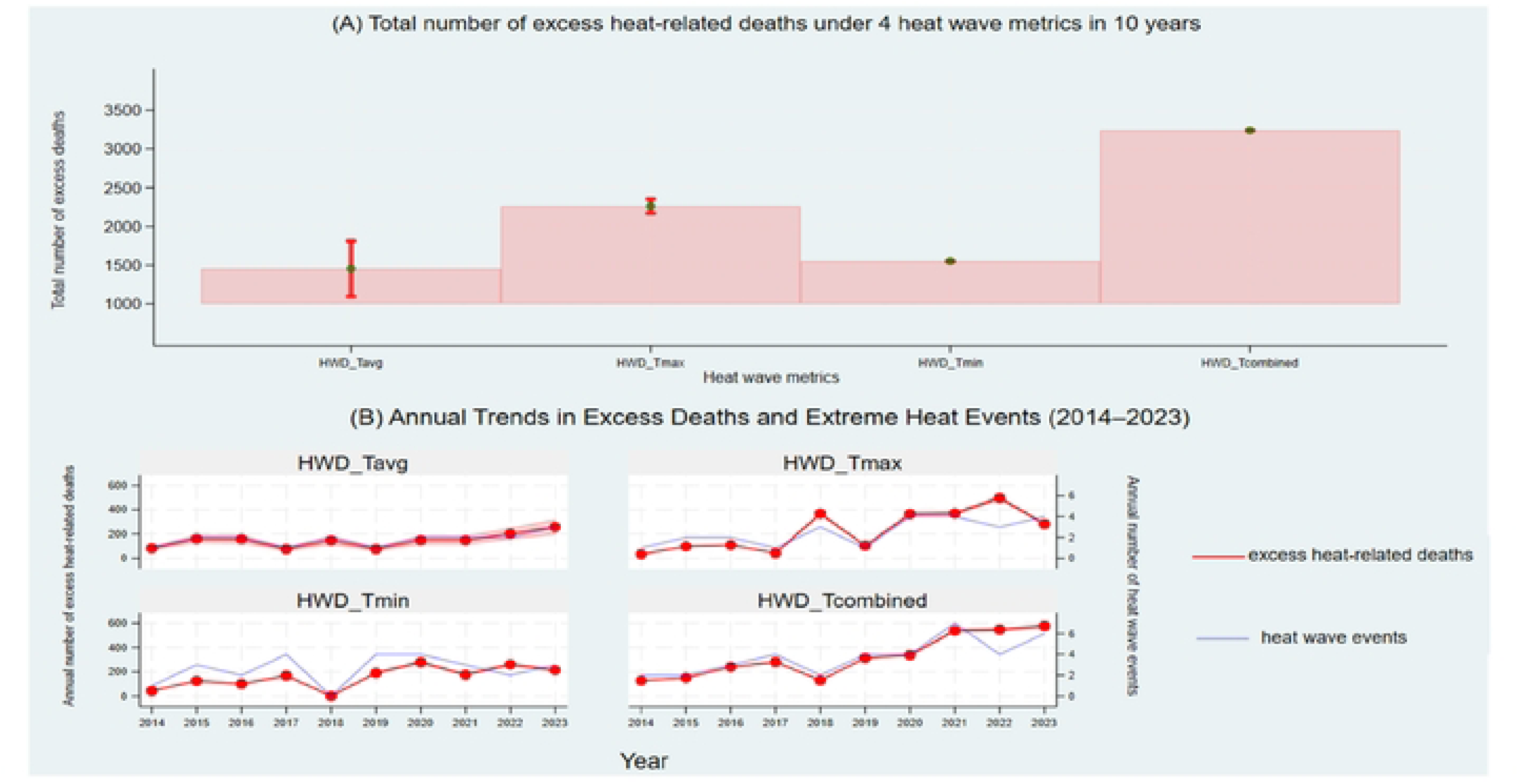
Excess heat-related deaths (2014–2023) under 4 definitions. A: Total number of excess heat-related deaths across 10 years; B: Annual number of excess heat-related deaths. Notes In Fig2A, the vertical lines extending from the top of each bar indicate the 95% CIs derived from Monte Carlo simulation. In Fig2B, the shaded areas around the lines indicate the 95% Cis derived from Monte Carlo simulation, providing a measure of the statical uncertainty around the annual excess death estimates.

**Table 2.**
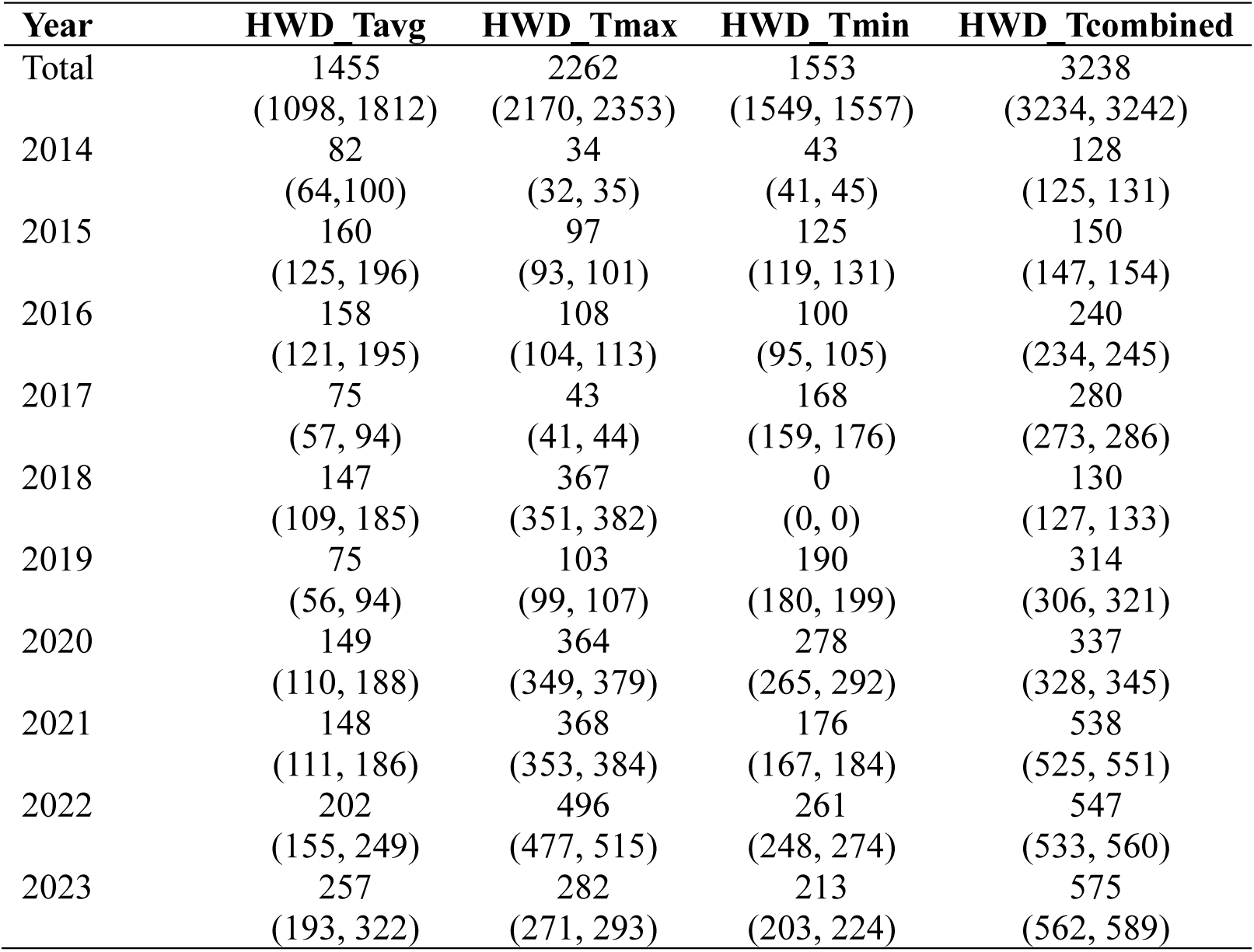
Cumulative excess deaths attributable to heat waves over the 10-year period under different heat wave definitions.

| **Year** | **HWD_Tavg** | **HWD_Tmax** | **HWD_Tmin** | **HWD_Tcombined** |
| --- | --- | --- | --- | --- |
| Total | 1455<br>(1098, 1812) | 2262<br>(2170, 2353) | 1553<br>(1549, 1557) | 3238<br>(3234, 3242) |
| 2014 | 82<br>(64,100) | 34<br>(32, 35) | 43<br>(41, 45) | 128<br>(125, 131) |
| 2015 | 160<br>(125, 196) | 97<br>(93, 101) | 125<br>(119, 131) | 150<br>(147, 154) |
| 2016 | 158<br>(121, 195) | 108<br>(104, 113) | 100<br>(95, 105) | 240<br>(234, 245) |
| 2017 | 75<br>(57, 94) | 43<br>(41, 44) | 168<br>(159, 176) | 280<br>(273, 286) |
| 2018 | 147<br>(109, 185) | 367<br>(351, 382) | 0<br>(0, 0) | 130<br>(127, 133) |
| 2019 | 75<br>(56, 94) | 103<br>(99, 107) | 190<br>(180, 199) | 314<br>(306, 321) |
| 2020 | 149<br>(110, 188) | 364<br>(349, 379) | 278<br>(265, 292) | 337<br>(328, 345) |
| 2021 | 148<br>(111, 186) | 368<br>(353, 384) | 176<br>(167, 184) | 538<br>(525, 551) |
| 2022 | 202<br>(155, 249) | 496<br>(477, 515) | 261<br>(248, 274) | 547<br>(533, 560) |
| 2023 | 257<br>(193, 322) | 282<br>(271, 293) | 213<br>(203, 224) | 575<br>(562, 589) |

**Figure 3** presents the annual trends in excess death rates (per 100,000 individuals) in Hong Kong from 2014 to 2023 under various heat wave definitions and compares these rates to Hong Kong’s age adjusted diabetes mortality. Under the HWD_Tmax and HWD_Tcombined definitions, the excess death rate attributable to heat waves exceeded that of diabetes in nearly half of the study period (from 2020 to 2023). Notably, under the HWD_Tcombined definition, the excess death rate was nearly twice that of diabetes in the recent three years (**Table 3**). Consequently, heat waves exhibiting high-risk patterns are poised to become the tenth leading cause of death in Hong Kong, even surpassing diabetes.^31^

**Figure 3.**
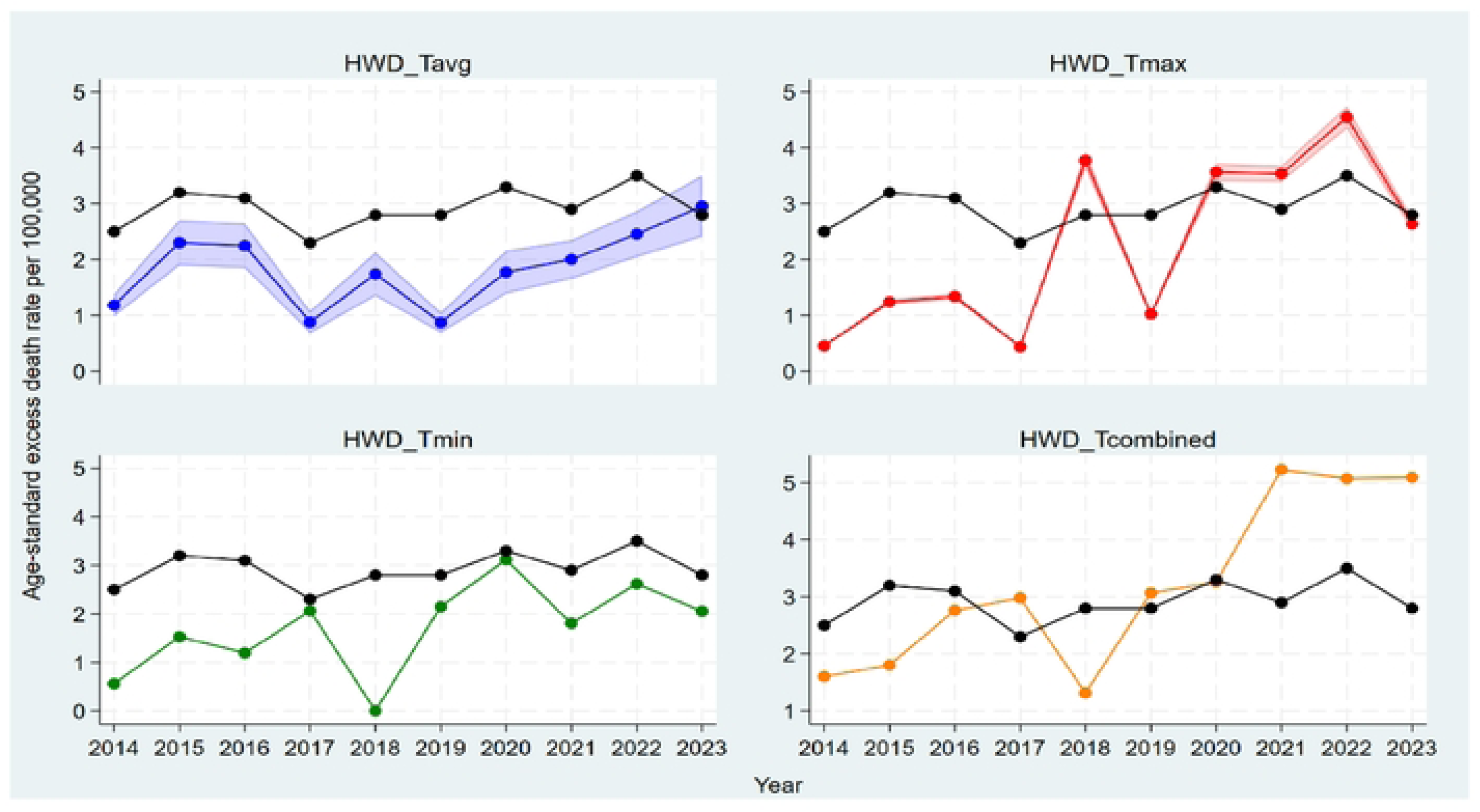
Annual Trends in Age-Standardized Excess Death Rates in Hong Kong (2014-2023) Note: The black line depicts the age-standardized mortality rate for diabetes in Hong Kong, serving as a comparative benchmark for heat wave-related excess deaths.

**Table 3.**
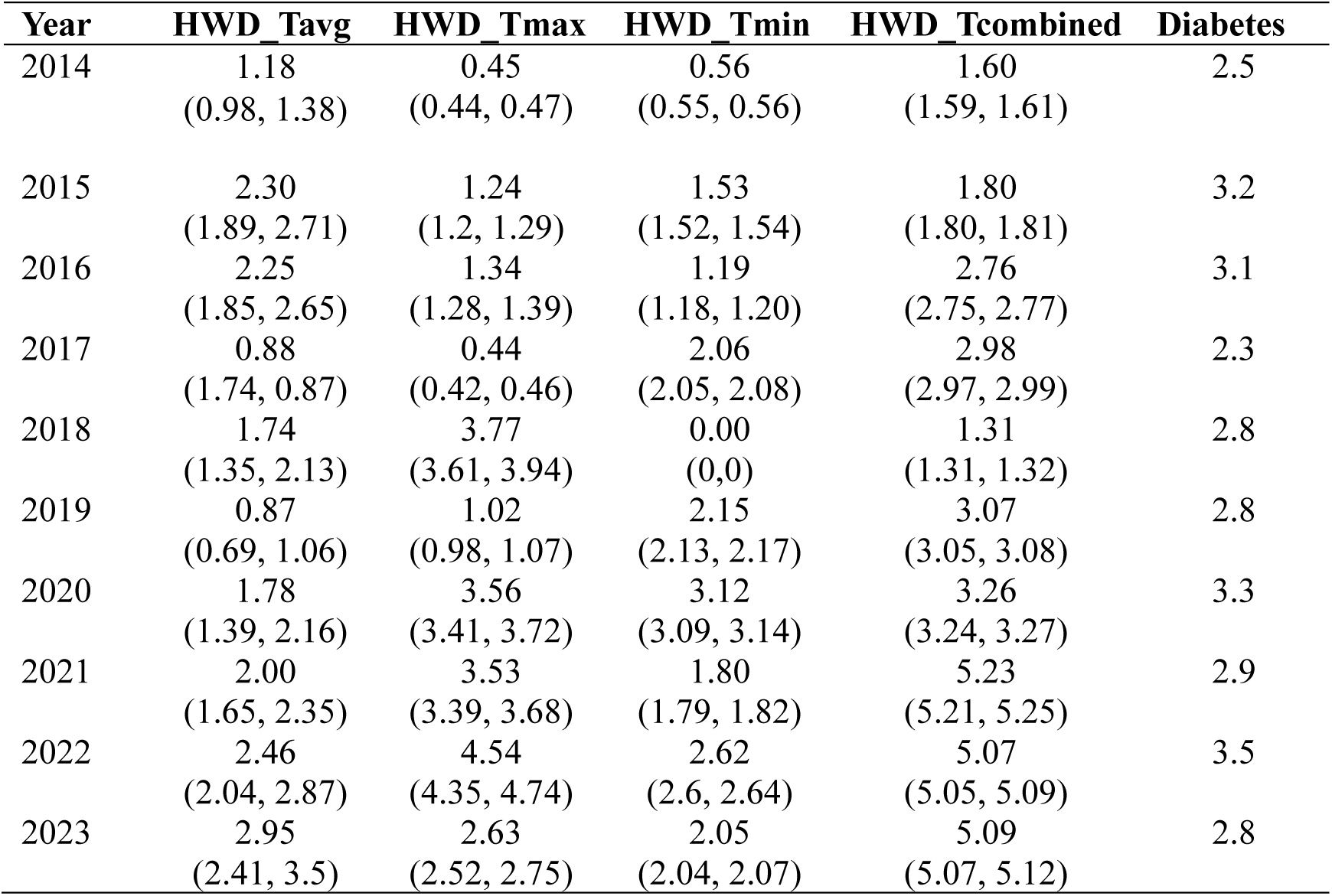
Annual Age-standardized excess death Rate per 100,000 population in the years 2014-2023 period.

| **Year** | **HWD_Tavg** | **HWD_Tmax** | **HWD_Tmin** | **HWD_Tcombined** | **Diabetes** |
| --- | --- | --- | --- | --- | --- |
| 2014 | 1.18<br>(0.98, 1.38) | 0.45<br>(0.44, 0.47) | 0.56<br>(0.55, 0.56) | 1.60<br>(1.59, 1.61) | 2.5 |
| 2015 | 2.30<br>(1.89, 2.71) | 1.24<br>(1.2, 1.29) | 1.53<br>(1.52, 1.54) | 1.80<br>(1.80, 1.81) | 3.2 |
| 2016 | 2.25<br>(1.85, 2.65) | 1.34<br>(1.28, 1.39) | 1.19<br>(1.18, 1.20) | 2.76<br>(2.75, 2.77) | 3.1 |
| 2017 | 0.88<br>(1.74, 0.87) | 0.44<br>(0.42, 0.46) | 2.06<br>(2.05, 2.08) | 2.98<br>(2.97, 2.99) | 2.3 |
| 2018 | 1.74<br>(1.35, 2.13) | 3.77<br>(3.61, 3.94) | 0.00<br>(0,0) | 1.31<br>(1.31, 1.32) | 2.8 |
| 2019 | 0.87<br>(0.69, 1.06) | 1.02<br>(0.98, 1.07) | 2.15<br>(2.13, 2.17) | 3.07<br>(3.05, 3.08) | 2.8 |
| 2020 | 1.78<br>(1.39, 2.16) | 3.56<br>(3.41, 3.72) | 3.12<br>(3.09, 3.14) | 3.26<br>(3.24, 3.27) | 3.3 |
| 2021 | 2.00<br>(1.65, 2.35) | 3.53<br>(3.39, 3.68) | 1.80<br>(1.79, 1.82) | 5.23<br>(5.21, 5.25) | 2.9 |
| 2022 | 2.46<br>(2.04, 2.87) | 4.54<br>(4.35, 4.74) | 2.62<br>(2.6, 2.64) | 5.07<br>(5.05, 5.09) | 3.5 |
| 2023 | 2.95<br>(2.41, 3.5) | 2.63<br>(2.52, 2.75) | 2.05<br>(2.04, 2.07) | 5.09<br>(5.07, 5.12) | 2.8 |

### Sub-group analysis

Our estimate of excess death counts by age and sex reflect the epidemiological variability from the original estimates of RRs (**Supplementary Table S3** and **Figure S4, S6**). Males experience nearly twice the number of excess deaths compared to females across most metrics, except for the nighttime heat definition (**Supplementary Figure S4A**). The magnitude of heat wave–related mortality risk increases with age, as individuals aged over 85 years appear to be the most vulnerable, with more than twice the number of deaths compared to other age groups across most metrics (**Supplementary Figure S4B**). Similar demographic variability is evident in the age-standardized excess death rates (**Supplementary Figure S5**).

## Discussion

Overall, our estimates of excess deaths in Hong Kong under different heat wave metrics ranged from 1455 to 3238 over a 10-year period. This finding helps to compare the magnitude of risk established in previous studies^10, 11, 12^ to benchmark conditions like diabetes. Notably, the magnitude of excess heat deaths shows variability across different heat wave metrics, highlighting the complexity of this public health issue.

Furthermore, our investigation reveals that extreme heat, if considered a distinct condition, would emerge as a leading cause of mortality in Hong Kong, with death rates nearly comparable to those of diabetes over the past decade. Specifically, the excess death rate ranged from 2.95 to 5.09 per 100,000 in 2023, positioning it among the top 10 causes of death in the region.^31^

Importantly, our findings reaffirm previous research indicating that individuals aged over 64 years in Hong Kong exhibit the highest susceptibility to heat wave-related excess mortality. This vulnerability is evidenced by both the excess number of deaths and the elevated death rates observed in this demographic. This well-known finding is attributed to a decreasing thermoregulatory capacity over time, as well as increased exposure to vasoactive medications for chronic diseases, which make the elderly more prone to health hazards during hot weather.^32, 33^

Behavioral change could prevent excess heat deaths. Some elderly individuals in Hong Kong have shown reluctance to use effective cooling appliances to save on energy costs, even if air conditioners are installed in their homes. ^34^ Many flats still have poor ventilation design and small window sizes characteristic of residential buildings in Hong Kong.^35^ Therefore, tailored protective measures based on population vulnerability and the specific characteristics of heat waves could prevent excess deaths. Clinicians including pharmacists, nurses, and physicians can be more proactive in assessing and counseling patients about their risk from age, co-morbidities, and medications. Social agencies conducting home visits can also engage in solution finding that is in harmony with cultural beliefs.

### Strengths and Policy Implications

To the best of our knowledge, this study is the first to estimate excess heat-related mortality in Hong Kong, encompassing both the direct and indirect impacts of extreme heat. While previous research has primarily focused on the relationship between temperature and mortality through relative risk estimates, our study shifts the emphasis to the absolute number of excess deaths—a more tangible and actionable metrics for policymakers and the public. Using the best available local evidence, we generated these estimates while accounting for a range of scenarios to reflect the complex and multifaceted nature of heat exposure and its health impacts.

Under global warming, Hong Kong is projected to experience more hot days and fewer cold days in the future,^36^ making it imperative for policymakers to develop adaptation strategies to protect vulnerable populations, especially in the context of the region’s rapidly aging population. In response to such challenges, many countries have introduced Heat Action Plans (HAPs), which include early warning systems, public advisories, and emergency health measures to reduce heat-related morbidity and mortality.^37^ Empirical evidence indicates that Heat Action Plans (HAPs) can effectively mitigate the impact of extreme temperatures. For instance, studies in Ahmedabad and France reported significant reductions in all-cause mortality following HAP implementation, with Ahmedabad avoiding 2,380 deaths on heat alert days and France reporting 4,388 fewer deaths during the 2006 European Heatwave. ^38, 39^ Similarly, a study in Montreal showed a reduction in daily mortality post-HAP, particularly among the elderly and lower socioeconomic status groups, thereby addressing heat-related health inequities. ^40^ Despite these successes, Hong Kong lacks a specific HAP tailored to its vulnerable population, highlighting the urgent need for an integrated heat-response strategy within the health and social care system.

### Limitations

Using local evidence can be a double-edged sword. First, in the absence of a universally accepted standard for measuring human exposure to heat, our study provides four distinct estimates of excess heat-related deaths, each based on different definitions of heatwaves used in previous local studies.^9, 11^ These measures can vary significantly depending on how heatwaves are characterized, such as by average temperature,^9^ maximum temperature, or nighttime temperature, and how deaths occurring in the days or weeks following a heatwave are accounted for.^11^ Consequently, there is no single, universally accepted method for attributing deaths to heat, resulting in a range of estimates rather than a definitive value. While it remains challenging to determine which estimates are the best, we offer these as a range, inviting readers to consider the broad spectrum of possible outcomes. Second, we assumed the entire territory of Hong Kong population is at equal risk of heat exposure, but heat levels vary spatially, and risk levels vary by education, occupation, socioeconomic status, and physical condition. Further stratification studies are needed to explore demographic heterogeneity.

## Conclusion

This study estimated the excess death count attributable to heat in Hong Kong over the past decade and leverages the best available local evidence to provide a range of scenarios, each grounded in methodologies established by previous research. Notably, individuals aged over 65 years were disproportionately affected, underscoring the heightened vulnerability of the senior population to extreme heat.

Because heat deaths are theoretically preventable, our study highlights the critical importance of developing territory-wide HAPs. Cities that convene multiple branches of government and civil society to tailor a local culturally appropriate approach have demonstrated success in controlling preventable deaths. Without such measures, extreme heat will continue to grow as a major cause of mortality in Hong Kong, posing significant public health challenges in the context of ongoing climate change.

## Data Availability

All relevant data are within the manuscript and its Supporting Information files.

## Author Contributions

Concept or design: DMB

Acquisition of data: JL, CR

Analysis or interpretation of data: ZL

Drafting of the manuscript: ZL, YK

Critical revision of the manuscript for important intellectual content: CR, JL, DMB.

All data used in this paper is in the public domain and is listed in the citations. CHATGPT-4o was used in the writing process to improve readability and language of the work, but not to analyze or interpret data or draw scientific conclusions.

## Declaration

The results of this study were presented in the Hong Kong College of Community Medicine Annual Scientific Meeting in Hong Kong (28 September 2024).

## Ethics approval

This study did not require ethical approval as it exclusively used publicly available data obtained from previously published. No new data was collected from human participants, and all analyses were conducted on anonymized, aggregated datasets from public sources.

## Supplementary Materials

**Supplementary Table S1.**
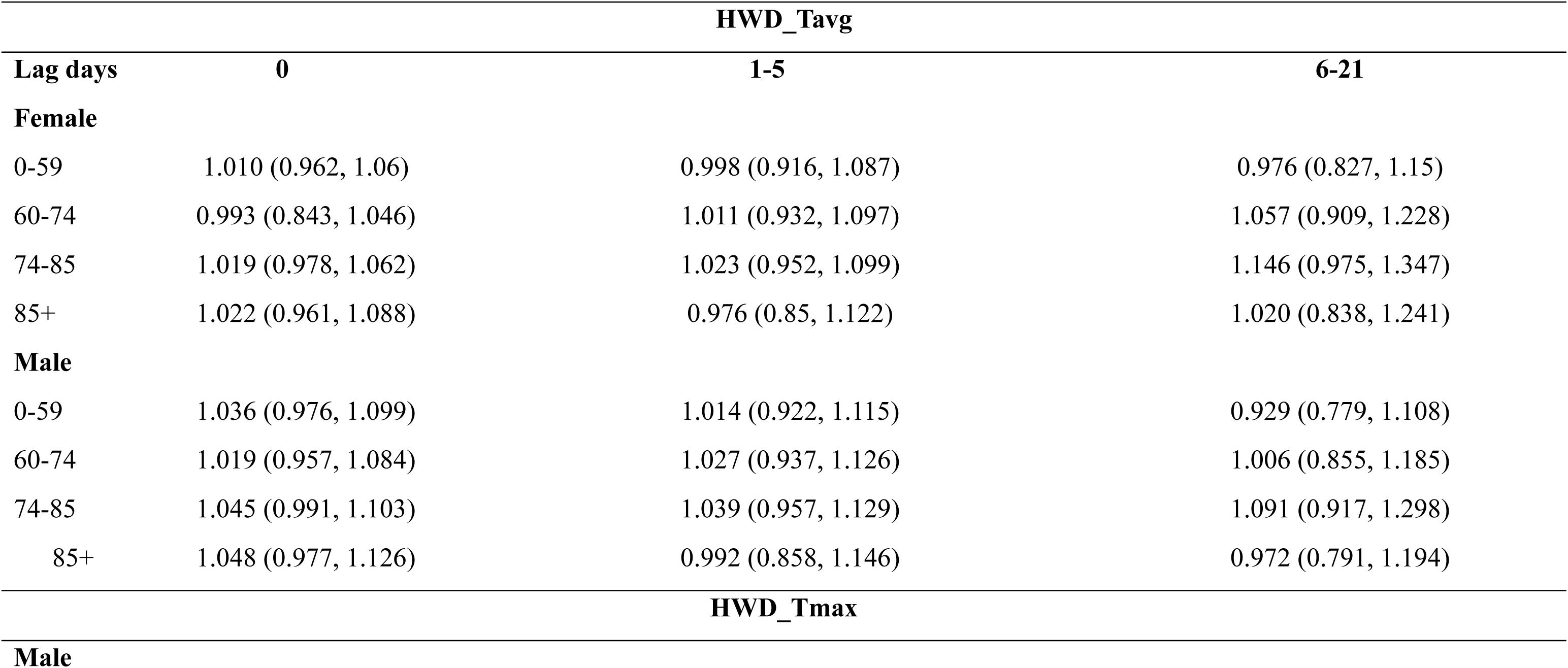
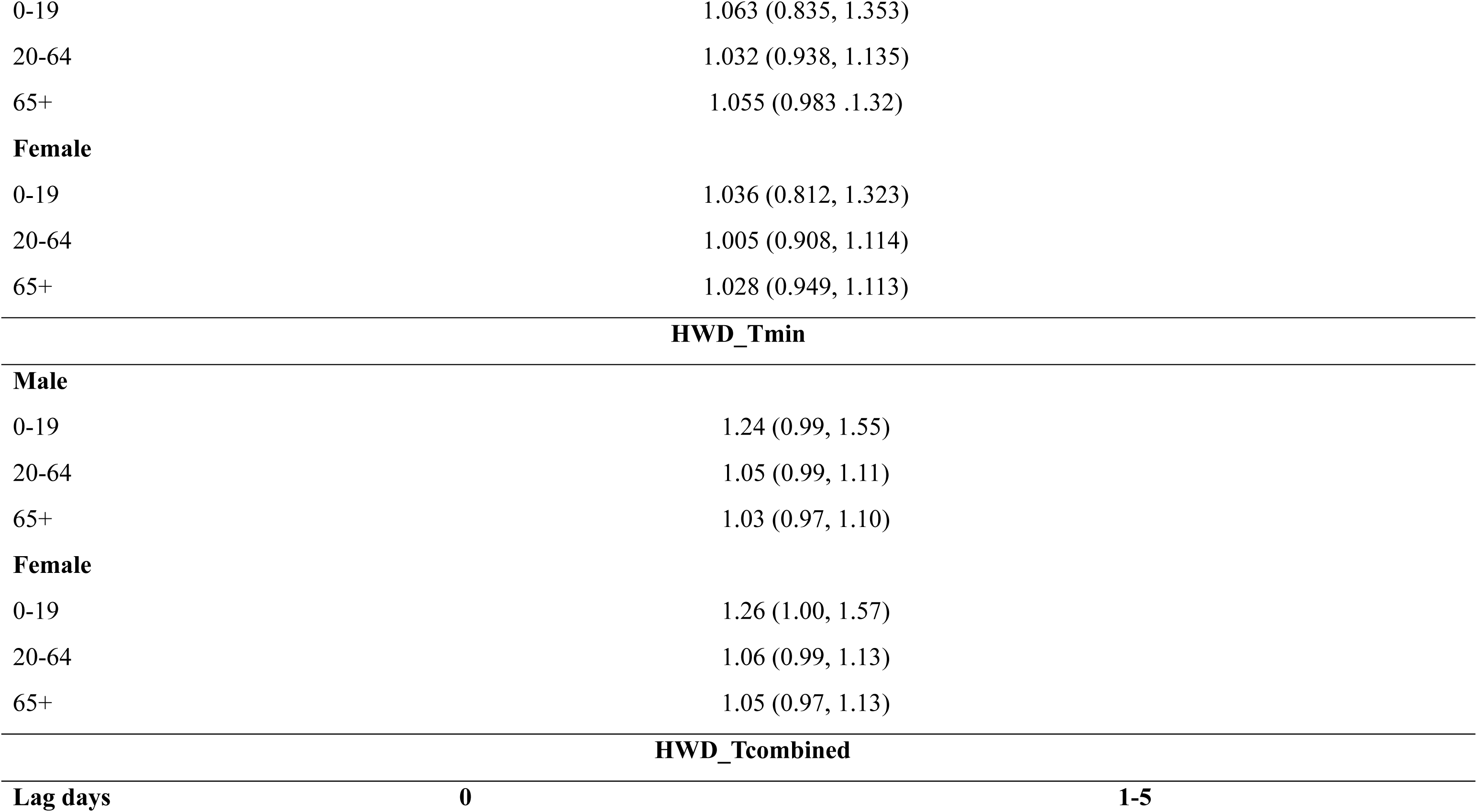
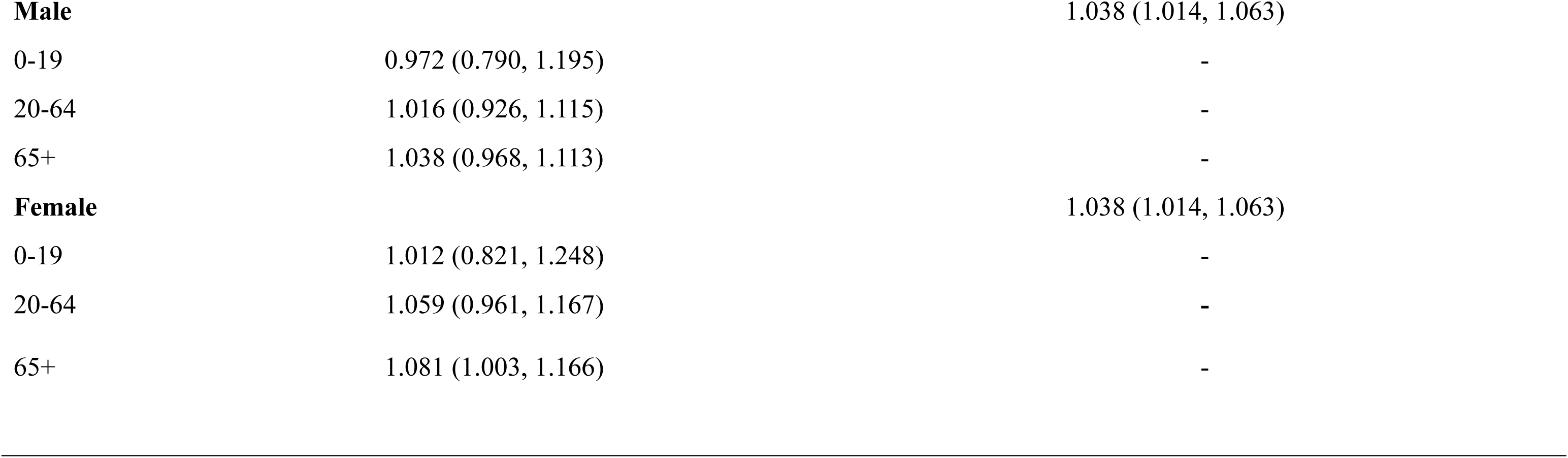
Age-sex specific Relative risk of extreme hot temperature on mortality and sub-categories over multiple lag days in Hong Kong: under four different heat wave definitions.

| Lag days | HWD_Tavg |  |  |
| --- | --- | --- | --- |
|  | 0 | 1-5 | 6-21 |
| **Female** |  |  |  |
| 0-59 | 1.010 (0.962, 1.06) | 0.998 (0.916, 1.087) | 0.976 (0.827, 1.15) |
| 60-74 | 0.993 (0.843, 1.046) | 1.011 (0.932, 1.097) | 1.057 (0.909, 1.228) |
| 74-85 | 1.019 (0.978, 1.062) | 1.023 (0.952, 1.099) | 1.146 (0.975, 1.347) |
| 85+ | 1.022 (0.961, 1.088) | 0.976 (0.85, 1.122) | 1.020 (0.838, 1.241) |
| **Male** |  |  |  |
| 0-59 | 1.036 (0.976, 1.099) | 1.014 (0.922, 1.115) | 0.929 (0.779, 1.108) |
| 60-74 | 1.019 (0.957, 1.084) | 1.027 (0.937, 1.126) | 1.006 (0.855, 1.185) |
| 74-85 | 1.045 (0.991, 1.103) | 1.039 (0.957, 1.129) | 1.091 (0.917, 1.298) |
| 85+ | 1.048 (0.977, 1.126) | 0.992 (0.858, 1.146) | 0.972 (0.791, 1.194) |
| **HWD_Tmax** |  |  |  |
| **Male** |  |  |  |

|  |  |
| --- | --- |
| 0-19 | 1.063 (0.835, 1.353) |
| 20-64 | 1.032 (0.938, 1.135) |
| 65+ | 1.055 (0.983 .1.32) |

**Female**
|  |  |
| --- | --- |
| 0-19 | 1.036 (0.812, 1.323) |
| 20-64 | 1.005 (0.908, 1.114) |
| 65+ | 1.028 (0.949, 1.113) |

**Male**
|  |  |
| --- | --- |
| 0-19 | 1.24 (0.99, 1.55) |
| 20-64 | 1.05 (0.99, 1.11) |
| 65+ | 1.03 (0.97, 1.10) |

**Female**
|  |  |
| --- | --- |
| 0-19 | 1.26 (1.00, 1.57) |
| 20-64 | 1.06 (0.99, 1.13) |
| 65+ | 1.05 (0.97, 1.13) |

|  |  |  |
| --- | --- | --- |
| **Male** |  | 1.038 (1.014, 1.063) |
| 0-19 | 0.972 (0.790, 1.195) | - |
| 20-64 | 1.016 (0.926, 1.115) | - |
| 65+ | 1.038 (0.968, 1.113) | - |
| **Female** |  | 1.038 (1.014, 1.063) |
| 0-19 | 1.012 (0.821, 1.248) | - |
| 20-64 | 1.059 (0.961, 1.167) | - |
| 65+ | 1.081 (1.003, 1.166) | - |

**Supplementary Table S2.**
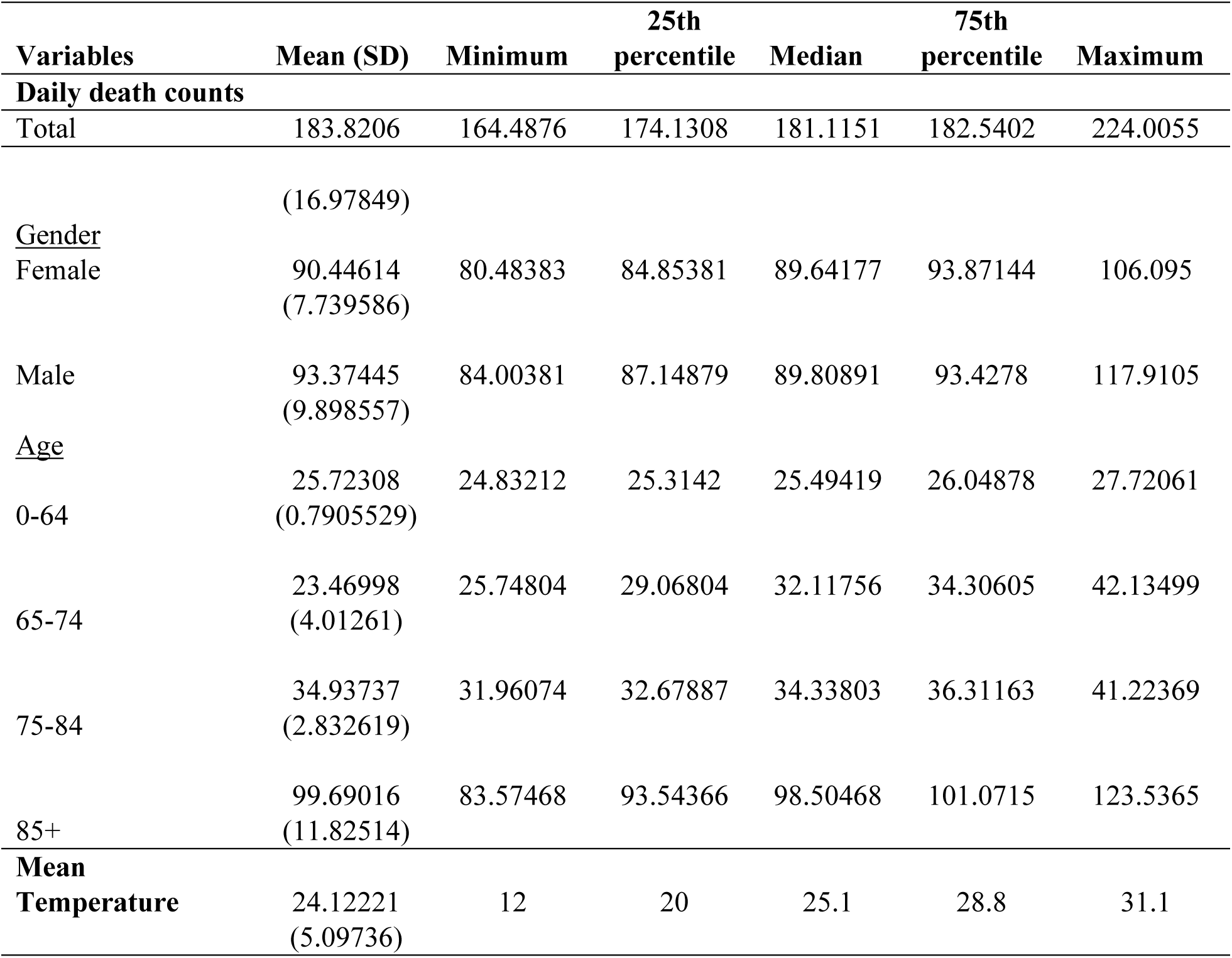
Descriptive statistics of daily deaths, mean temperature.

| **Variables** | **Mean (SD)** | **Minimum** | **25th percentile** | **Median** | **75th percentile** | **Maximum** |
| --- | --- | --- | --- | --- | --- | --- |
| **Daily death counts** |  |  |  |  |  |  |
| Total | 183.8206 | 164.4876 | 174.1308 | 181.1151 | 182.5402 | 224.0055 |
|  | (16.97849) |  |  |  |  |  |
| <u>Gender</u> |  |  |  |  |  |  |
| Female | 90.44614<br>(7.739586) | 80.48383 | 84.85381 | 89.64177 | 93.87144 | 106.095 |
| Male | 93.37445<br>(9.898557) | 84.00381 | 87.14879 | 89.80891 | 93.4278 | 117.9105 |
| <u>Age</u> |  |  |  |  |  |  |
| 0-64 | 25.72308<br>(0.7905529) | 24.83212 | 25.3142 | 25.49419 | 26.04878 | 27.72061 |
| 65-74 | 23.46998<br>(4.01261) | 25.74804 | 29.06804 | 32.11756 | 34.30605 | 42.13499 |
| 75-84 | 34.93737<br>(2.832619) | 31.96074 | 32.67887 | 34.33803 | 36.31163 | 41.22369 |
| 85+ | 99.69016<br>(11.82514) | 83.57468 | 93.54366 | 98.50468 | 101.0715 | 123.5365 |
| **Mean Temperature** | 24.12221<br>(5.09736) | 12 | 20 | 25.1 | 28.8 | 31.1 |

**Supplementary Table S3.** Demographic heterogeneity in total number of excess heat-related deaths: by age and sex groups.

|  | **HWD_Tavg** | **HWD_Tmax** | **HWD_Tmin** | **HWD_Tcombined** |
| --- | --- | --- | --- | --- |
| **Sex** |  |  |  |  |
| Female | -11 (-201, 178) | 802 (755, 849) | 877 (837, 918) | 1154 (1117, 1192) |
| Male | 1466 (1294, 1638) | 1459 (1413, 1505) | 676 (639, 713) | 2083 (2042, 2124) |
| **Age** |  |  |  |  |
| 0-64 | -45 (-88, -3) | 241 (224, 258) | 281 (270, 291) | 382 (368, 397) |
| 65-74 | 320 (281, 360) | 342 (331, 353) | 183 (172, 193) | 481 (471, 491) |
| 75-84 | 1331 (1269, 1393) | 465 (449, 480) | 263 (249, 276) | 656 (643, 669) |
| 85+ | -152 (-366, 63) | 1214 (1167, 1260) | 828 (784, 871) | 1719 (1679, 1759) |

**Supplementary Figure 4.**
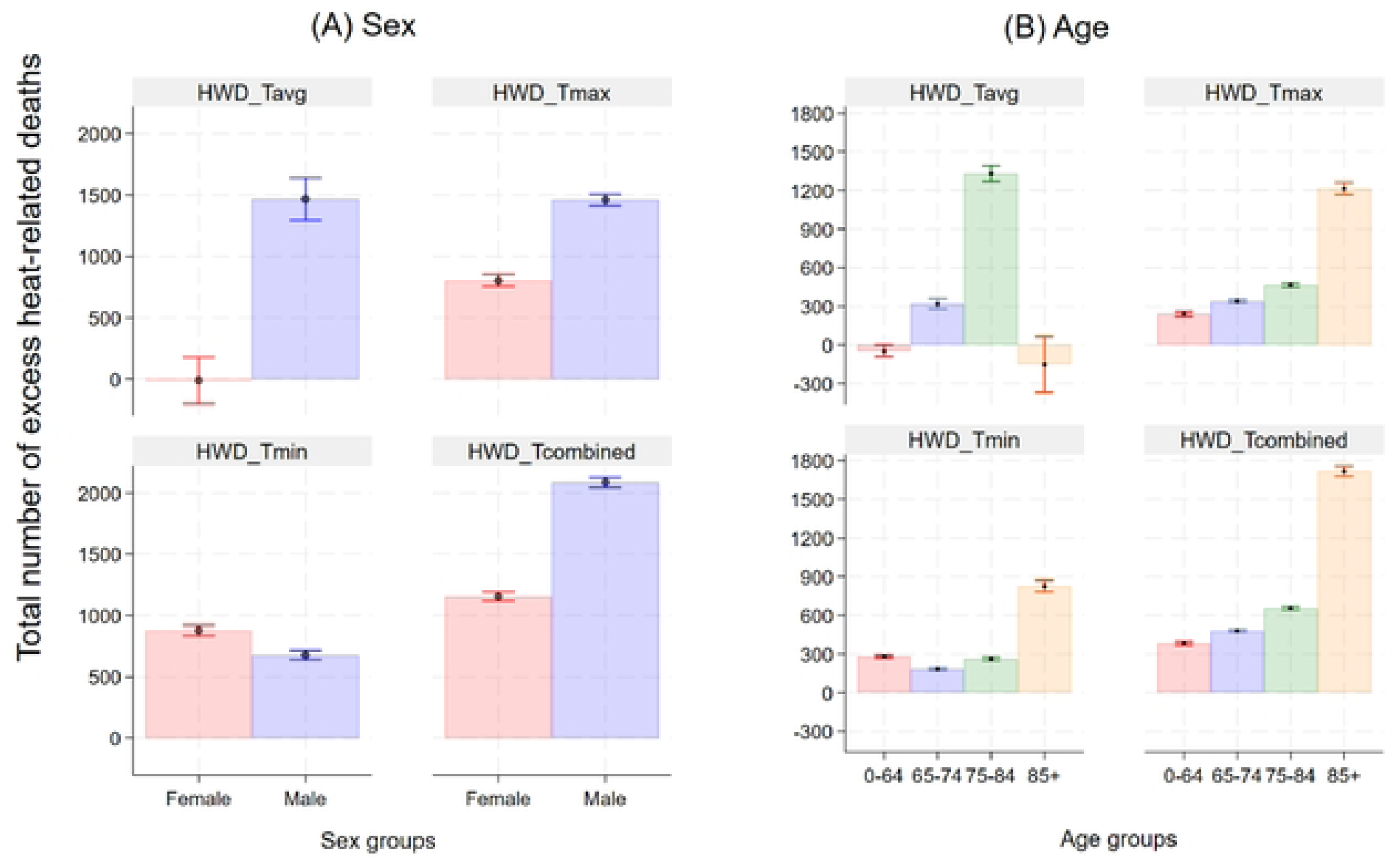
Demographic heterogeneity in total number of excess heat-related deaths: by age and sex groups.

**Supplementary Figure 5.**
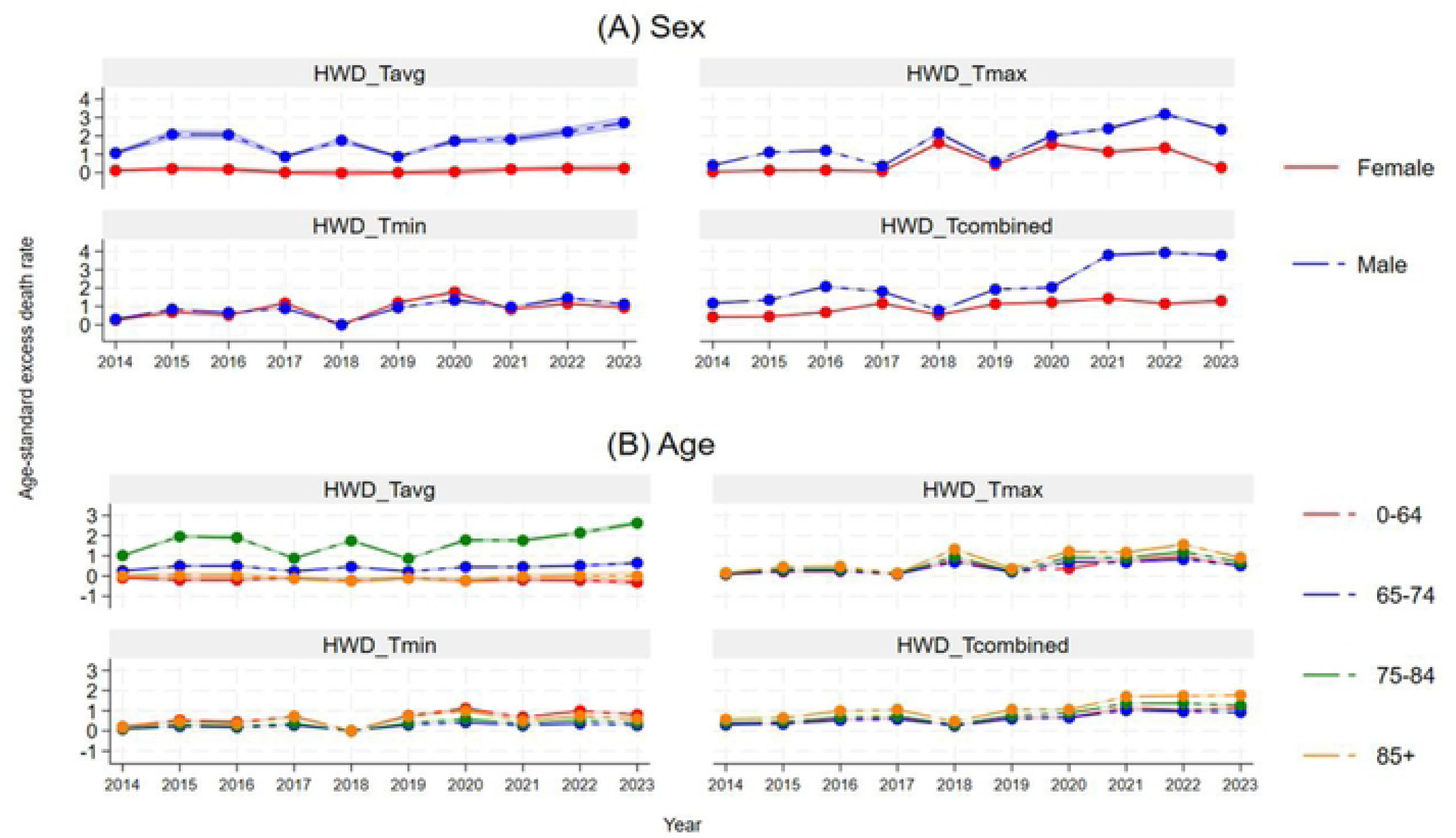
Demographic Heterogeneity in age-standardized mortality rates: by age and sex groups.

